# Post-COVID-19 tele-survey for persistent symptoms in a single center hospital cohort in India along with a parallel country-wide web-survey

**DOI:** 10.1101/2022.02.20.22271119

**Authors:** Kausik Chaudhuri, Jit Sarkar, Tirthankar Das, Shekhar Ranjan Paul, Rajita Basu, Supratik Gangopadhyay, Devyani Gangopadhyay, Asis Manna, Anima Haldar, Yogiraj Ray, Sayantan Banerjee, Dipyaman Ganguly

## Abstract

**Introduction:** A major concern amidst the ongoing coronavirus pandemic has been the longer term persistence of morbidities in individuals recovering from COVID-19 disease, called ‘long COVID’. We aimed at documenting the prevalence and key associations of post-COVID symptoms (PCS) in India in telephonic survey among recovered patients in a single hospital in eastern India as well as a parallel web-survey covering a wider population of the country.

**Methods:** Self-reported PCS, ranging up to one year since the original COVID-19 diagnosis, were documented in a telephonic survey of subjects (analyzed N=986), treated for acute COVID-19 in Infectious Diseases and Beleghata General Hospital, Kolkata, between April 1, 2020 and April 13, 2021. In parallel, we ran a web-based survey (analyzed N=580), to evaluate concordance.

**Results:** Shortness of breath, fatigue and insomnia were identified to be the most commonly reported PCS in both the surveys, with higher prevalence in females. In the telephonic survey, a 3.65% post-discharge mortality was registered within a median of 39 days since COVID diagnosis. Intensive care during acute disease and hypertension were more often associated with PCS, while fatigue was more often reported by the 20-40 years age-group. The web-survey revealed a gradual decline in PCS with time since COVID-19 diagnosis and type 2 diabetes to be associated with higher prevalence of these symptoms.

**Conclusions:** We assessed the predominant PCS among Indian COVID-19 patients and identified key demographic and clinical associations in our surveys, which warrants deeper epidemiological and mechanistic studies for guiding management of long-COVID in the country.

## Introduction

The coronavirus pandemic due to SARS-CoV-2 outbreak has driven considerable infections and mortality across all the populated continents (1,2). A systemic hyper-inflammatory surge has been associated with the severe COVID-19 (3-5). A major concern amidst this pandemic has been the long term morbidities in the individuals recovering from COVID-19 disease, the so-called ‘long COVID’ (6,7). The ‘long-haul COVID’, which has emerged as a definite clinical entity all over the world, has largely been attributed to the undue lingering of the systemic inflammation for variable time periods (8). Exploring the factors that make convalescent subjects more susceptible to long-COVID is a domain of active research. A considerable number of prospective follow-up studies and telephone surveys done in different parts of the world have been asserting about the commonly reported post-COVID-symptoms (PCS) and revealing the heterogeneity of susceptibility among different cohorts, based on ethnicity, age-groups, pre-existing co-morbidities as well as nature of the acute disease (9-15).

In the present study we aimed at documenting self-reported PCS, ranging up to one year since the original COVID-19 diagnosis in a telephonic survey of subjects (analyzed N=986) who were treated for acute COVID-19 disease in a single hospital in eastern India, Infectious Diseases and Beleghata General (ID & BG) Hospital, Kolkata, between April 1, 2020 and April 13, 2021, to document perceived lingering symptoms. In parallel we ran a web-survey, which was responded by convalescent individuals from different parts of India (analyzed N=580), to evaluate concordance.

## Methods

Both the telephone survey for the hospital cohort and the web-survey were approved by the Institutional Ethics Committee of ID & BG Hospital (No. IDBGH/Ethics/4161). Verbal informed consents were taken in the telephonic survey. The introductory note on study information for the web survey informed the participants that their participation asserted informed consent for the study. Anonymized data were processed for analyses. Strength of association if any was validated using chi-square test, considering a p-value of 0.05 to be statistically significant.

## Results and Discussion

In the telephonic survey a total of 1040 individuals, who got discharged after COVID-19 disease remission from ID & BG hospital, Kolkata, were contacted. 38 post-remission deaths were recorded after disease remission (3.65%, close to 1 in 27 discharged patients, median time of post-remission death since COVID-19 diagnosis being 39.5 days with range of 2-398 days). Among them 13 were females (mean age 61.23 years) and 25 males (mean age 69 years).

Data on PCS from 1002 individuals were collected, out of which data from 986 individuals could be analyzed. The subjects were largely located in the Kolkata Metropolitan Area, in the eastern Indian state of West Bengal (Figure 1A). Among the clinical symptoms about which self-reporting was sought for actively, three major symptoms were shortness of breath (SOB, 11.43%), fatigue (13.54%) and insomnia (7.02%) (Figure 1B). Further subclass analyses were performed on the prevalence of these three. In the sub-class analyses association of these three PCS were evaluated with sex, age-groups, time elapsed since the COVID-19 diagnosis, requirement of intensive care during COVID-19 and two major co-morbidities, viz. type 2 diabetes mellitus (T2DM) and hypertension (HTN).

**Figure 1.**
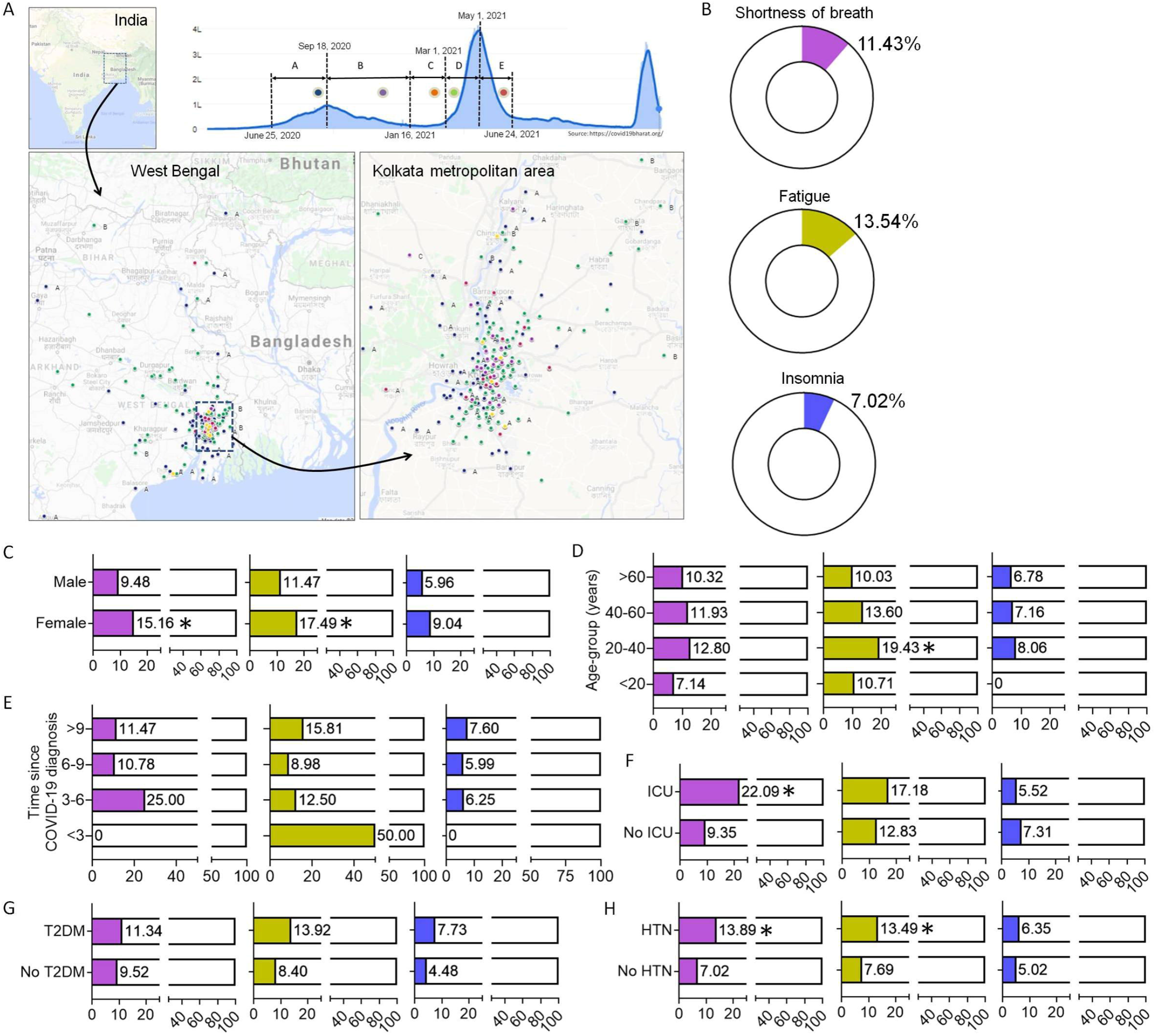
Documenting post-COVID symptoms in a telephonic survey in a single hospital cohort in eastern India. **A)** Mapping the residential origins of the subjects participating the telephonic survey with color-coding for five time-periods of the epidemic when they got infected with SARS-CoV-2. **B)** Prevalence of three major post-COVID symptoms among all subjects. The symptoms are color-coded for the rest of the figure. **C)** Comparing prevalence of post-COVID symptoms between females and males. **D)** Comparing prevalence of post-COVID symptoms among indicated age-groups. **E)** Comparing prevalence of post-COVID symptoms among subjects based on time since COVID-19 diagnosis. **F)** Comparing prevalence of post-COVID symptoms among subjects based on whether or not intensive care was required during acute COVID-19. **G)** Comparing prevalence of post-COVID symptoms between subjects with and without type 2 diabetes. **H)** Comparing prevalence of post-COVID symptoms between subjects with and without hypertension.

Self-reporting of post-COVID SOB symptoms was considerably higher among females (Figure 1C). 15.16% of females reporting it compared to 9.48% among males (p=0.01). Similarly complaints of post-COVID fatigue was higher among females (17.49%) compared to males (11.47%). Interestingly, among the age-groups compared subjects from the 20-40 years age-group reported post-COVID fatigue much more frequently (19.43%, p=0.02). For other two symptoms no significant age-group variation was noted (Figure 1D). The time of reporting since the COVID-19 diagnosis (stratified as < 3, 3-6, 6-9 and >9 months since diagnosis) was not associated with the analyzed PCS (Figure 1E). But of note here, the earlier time-period of <3 months was underrepresented among the telephone survey participants (N=3). Requirement of intensive care during the active COVID-19 was strongly associated with post-COVID SOB (22.09%, compared to 9.35% for patients not requiring intensive care, p=0.00) (Figure 1F). Higher propensity for post-COVID fatigue was also noted among patients requiring COVID-19 intensive care, although it was not statistically significant (17.18% vs 12.83%). Both T2DM and HTN was associated with higher propensity of post-COVID SOB and fatigue reported by the tele-survey subjects, association with HTN being statistically significant (Figure 1G & H).

To gather data from another geographically wider cohort from India, in parallel to the tele-survey we ran the web-form based survey. Anonymous data on PCS from 621 individuals were collected, out of which data from 580 individuals could be analyzed. As the web-form was in English and only available on the web the demographic representation was restricted to higher socio-economic strata urban population with a younger age distribution. Nevertheless, the web-survey widely represented India (analyzed from postal codes provided by the respondents) as shown in Figure 2A. The metropolitan cities of Kolkata in the eastern India, New Delhi in the northern India and Chennai in the southern India was over-represented. Again, the three major symptoms were SOB, fatigue and insomnia, but the frequency of reporting these PCS was much higher compared to the tele-survey (Figure 2B). This may represent a motivational bias among the symptomatic subjects to respond to the survey. In the web-survey the reporting of SOB was further categorised into SOB at rest, while walking and while climbing stairs.

**Figure 2.**
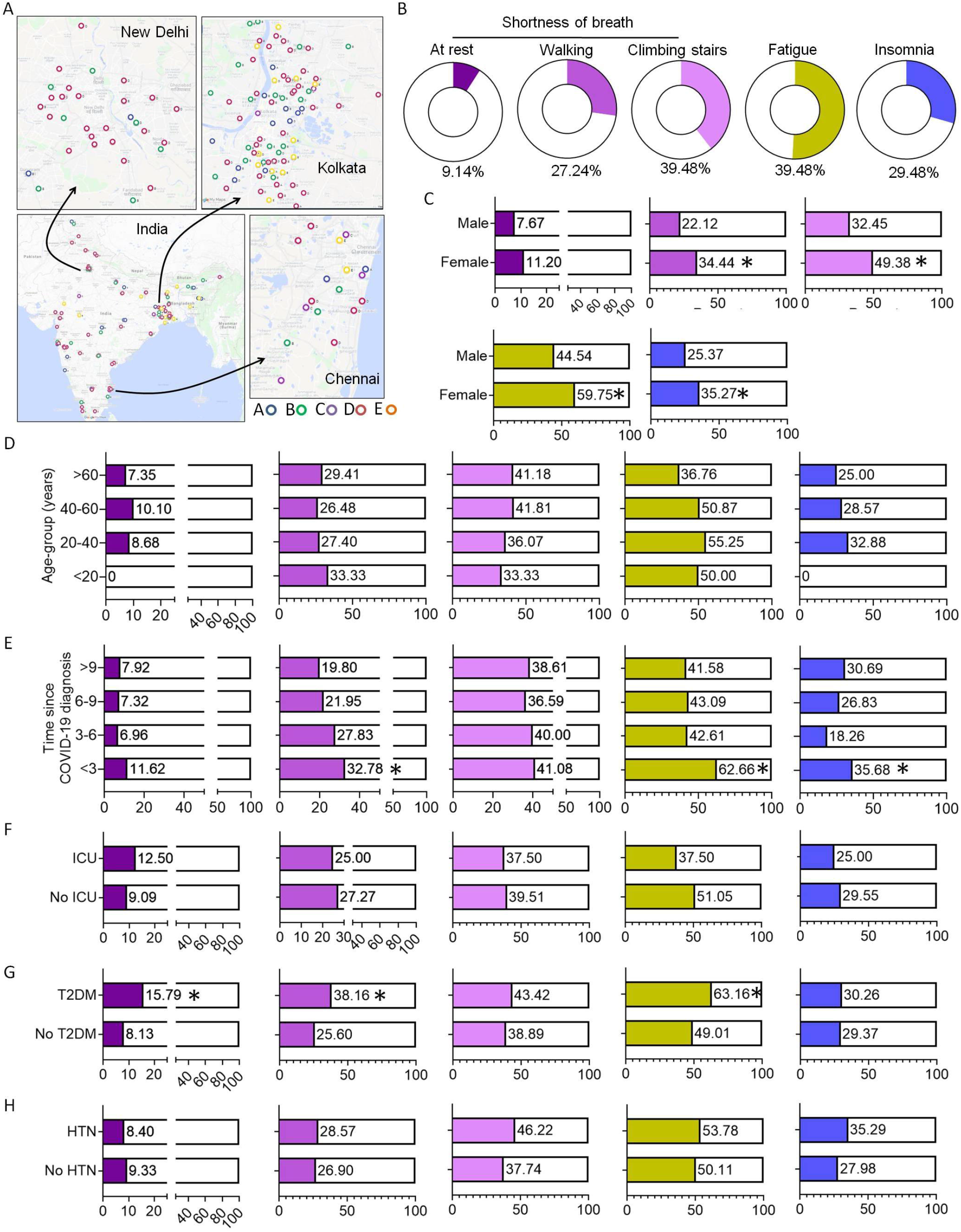
Documenting post-COVID symptoms in a web-survey in India. **A)** Mapping the residential origins of the subjects participating the web-survey with color-coding for five time-periods of the epidemic when they got infected with SARS-CoV-2, as shown in figure 1A. **B)** Prevalence of the major post-COVID symptoms among all subjects. The symptoms are color-coded for the rest of the figure. **C)** Comparing prevalence of post-COVID symptoms between females and males. **D)** Comparing prevalence of post-COVID symptoms among indicated age-groups. **E)** Comparing prevalence of post-COVID symptoms among subjects based on time since COVID-19 diagnosis. **F)** Comparing prevalence of post-COVID symptoms among subjects based on whether or not intensive care was required during acute COVID-19. **G)** Comparing prevalence of post-COVID symptoms between subjects with and without type 2 diabetes. **H)** Comparing prevalence of post-COVID symptoms between subjects with and without hypertension.

Self-reporting of post-COVID SOB symptoms while walking or climbing stairs as well as fatigue and insomnia was found to be significantly higher among (Figure 2C). No significant age-group variation was noted for the PCS (Figure 2D). The earlier time of reporting at <3 months since COVID-19 diagnosis was found to be significantly associated with the SOB while walking, fatigue as well as insomnia (Figure 2E), which was missed in the tele-survey perhaps due to under-representation of contacted subjects. On the other hand, requirement of intensive care (N=9) during acute COVID-19 was not associated with PCS in the web-survey, again perhaps due to the under-representation of such subjects (Figure 2F). Finally, both T2DM and HTN was associated with higher propensity of post-COVID SOB and fatigue reported by the web-survey subjects too, with the associations of T2DM with SOPB at rest and while walking as well as fatigue being statistically significant (Figure 2G & H).

## Discussion

The PCS found to be most prevalent in our study have also been reported by previous studies done in different parts of the world (6-20), although the relative distribution varied among cohorts, as did the demographic and clinical associations. The most extensive and long physical follow-up study, done in Wuhan, China, reported fatigue and muscle weakness (63%) and sleep difficulties (26%) to be the most prevalent PCS at a median of 186 days since symptom onset (N=1655) (9). This study also reported an increase in the prevalence of SOB from a median of 185 days to median of 348 days since symptom onset (N=1272) as well as a higher prevalence of PCS among females (10). Similarly in telephonic surveys done in UK (N=110) and France (N=120), beyond 3 months of symptom onset, fatigue was the most prevalent PCS (39% and 55% reporting it respectively), followed by SOB (39% and 41.7% respectively) and sleep disorder (24% and 30.8% respectively) (12, 14). In addition a large fraction of PCS was associated with intensive care for COVID-19 (16.4% and 20% respectively) in these studies.

Among the Indian studies, a prospective study done in a north Indian cohort of 1234 COVID-19 patients had reported 40.1% patients reporting persistent symptoms at clinical remission of the acute disease, while 9.9% had symptoms beyond 3 months since remission (15). This study also found SOB, fatigue and insomnia among the commonly reported lingering symptoms. In another Indian study a web-based survey had 2038 respondents with almost 40.1% reporting PCS beyond 3 months since the acute disease and fatigue was most commonly complained of (16). A north Indian single hospital study collected data from a younger cohort of patients (N=773, median age 34 years) and reported the prevalence of PCS (again most common being fatigue) to be 12.8% at 3 months since diagnosis. This study also reported PCS to be more prevalent in females (17).

Association of major co-morbidities like T2DM and HTN with the most prevalent PCS in our study is of great interest, especially with recent findings on prevalent post-COVID cardio-vascular morbidities discerned in a large cohort (20). Of note here, a smaller study in India with 160 patients reported SOB, fatigue and insomnia to be most prevalent PCS and them being associated with co-morbidities like T2DM and HTN (18).

Thus in this study we gathered valuable insights on the predominant PCS among Indian COVID-19 patients and identified key demographic and clinical associations through parallel single hospital telephonic survey and country-wide web-survey. These warrant deeper epidemiological and mechanistic studies for guiding management of long-COVID in the country.

## Data Availability

All data produced in the present study are available upon reasonable request to the authors.

## Contributors

KC, AM, AH, SB and DG designed the study; JS analyzed the data and performed statistics; TD helped with the telephonic survey and data curation; SRP and RB helped with telephonic survey data collection; YR helped with web-survey; SG and DG helped with geographic data analysis; KC, SB and DG conceptualized the study.

## Funding

The study was funded by P07 grant from CSIR-Indian Institute of Chemical Biology, Kolkata, to DG.

## Competing interest

Authors declare no competing interest.

## Notes

### Competing Interest Statement

The authors have declared no competing interest.

### Author Declarations

Ethics Committee of Infectious Disease and Beleghata General Hospital, Kolkata, India, gave ethical approval for this work.

